# An Interpretable Sparse Graph Contrastive Learning Approach for Identifying Breast Cancer Risk Variants

**DOI:** 10.1101/2025.01.13.25320451

**Authors:** Gudhe Naga Raju, Jaana M. Hartikainen, Maria Tengström, Katri Pylkäs, Robert Winqvist, Veli-Matti Kosma, Hamid Behravan, Arto Mannermaa

## Abstract

Genome-wide association studies (GWASs) have identified over 2,400 genetic variants associated to breast cancer. Conventional GWASs methods that analyze variants independently often overlook the complex genetic interactions underlying disease susceptibility. Machine and deep learning approaches present promising alternatives, yet encounter challenges, including overfitting due to high dimensionality (∼10 million variants) and limited sample sizes, as well as limited interpretability. Here, we present GenoGraph, a graph-based contrastive learning framework designed to address these limitations by modeling high-dimensional genetic data in low-sample-size scenarios. We demonstrate GenoGraph’s efficacy in breast cancer case-control classification task, achieving accuracy of 0.96 using the Biobank of Eastern Finland dataset. GenoGraph identified rs11672773 (*ZNF8*) as a key risk variant in Finnish population, with significant interactions with rs10759243 (*KLF4*) and rs3803662 (*TOX3*). Furthermore, in silico validation confirmed the biological relevance of these findings, underscoring GenoGraph’s potential to advance breast cancer risk prediction and elucidate genetic interactions for personalized medicine.

## INTRODUCTION

Single-nucleotide polymorphisms (SNPs) are the most prevalent form of genetic variants within the human genome. Each SNP represents a variation at a single nucleotide position, and a few serve as genetic markers in determining an individual susceptibility to breast cancer (BC).^1,2^ Approximately 5–10% of newly diagnosed BC cases are associated with familial genetic predisposition, underscoring the critical role of hereditary genetic factors in BC risk.^3^ Mutations in genes such as *BRCA1* and *BRCA2* significantly increase the lifetime risk of developing BC.^4^ Furthermore, mutations in high-risk genes such as *TP53, PTEN, CDH1, STK11,* and *PALB2* confer a 20–40% lifetime risk of BC.^5,6,7^ Interestingly, the prevalence of certain genetic mutations exhibits population-specific patterns. In the Finnish population, for instance, *PALB2* (*c.1592delT*) and *CHEK2* (*c.1100delC*) mutations are particularly prevalent, with frequencies of approximately 1% and 2.5%, respectively, and are associated with a 60–80% lifetime risk of BC. This emphasizes the importance of population-specific genetic studies in risk assessment and prevention strategies.^5,8^ These genetic markers also provide insights into the molecular mechanisms underlying the disease’s development.

Genome-wide association studies (GWASs) have been instrumental in identifying associations between genetic variants and diseases.^9,10^ To date, GWASs have identified 2,454 variants strongly associated with BC, including 209 variants linked to estrogen receptor (ER)-negative BC, 87 associated with ER-positive BC, 39 with progesterone receptor (PR)-negative BC, 2 with PR-positive BC, and 38 with triple-negative BC subtypes.^10^ A conventional GWAS relies on statistical approaches, such as the chi-square (𝜒²) test, to compute the 𝑝-values for genetic variants associated with specific traits or phenotypes.^9^ While these conventional approaches have provided valuable insights, they are limited in their ability to account for complex genetic interactions and correlations among multiple variants, although they are often crucial in understanding the genetic architecture of complex diseases like BC.^11,12^

Recently, machine learning (ML) and deep learning (DL) approaches have been employed in GWAS to capture more complex genetic interactions, which traditional methods may overlook.^11,12,13^ However, these approaches often suffer from the "curse of dimensionality," as they involve high-dimensional genetic data (≈10 million SNPs) with a limited sample size (HDLS). HDLS is an Nondeterministic Polynomial-time (NP)- hard problem and can lead to overfitting in ML and DL models, resulting in poor generalization and reduced predictive power.^14^ Feature selection (FS) techniques are often applied to mitigate this issue by identifying the most informative SNPs.^15^ Nguyen et al.^16^ developed a two-stage random forest (RF) method to filter informative SNPs, categorizing them into highly and weakly informative groups. Behravan et al.^11^ proposed an adaptive iterative SNP selection method to capture SNP-SNP interactions, improving BC risk prediction accuracy. Jo et al.^17^ introduced a DL-based sliding window association test (SWAT-CNN) to identify SNPs associated with Alzheimer’s disease. Recently, Li et al.^18^ developed a 14-layer deep neural network (DNN) model, named DeepGWAS, which outperformed traditional ML algorithms, including XGBoost and logistic regression, by integrating linkage disequilibrium and functional epigenomic data. Similarly, Arloth et al.^19^ demonstrated that integrating regulatory information into predictive models can further enhance the identification of genotype-phenotype associations, highlighting the importance of leveraging biologically relevant data to address the challenges of HDLS.

In this study, we introduce GenoGraph, a graph-based contrastive learning framework specifically designed to handle HDLS scenarios in genomics data. GenoGraph leverages a supervised contrastive learning approach^20,21^ to model the complex relationships between genetic variants, effectively reducing the risk of overfitting. The framework employs a sparse graph variational autoencoder (SGVAE) for contrastive learning to generate robust representations of genetic variants. Subsequently, integrates an interpretable graph explainer^22^ (GNNExplainer), which identifies and prioritizes the most predictive and interacting variants. By employing graph-based approach, GenoGraph enhances both the interpretability and reliability of variant embeddings, thus overcoming the shortcomings related to HDLS and variant interactions associated with traditional GWAS methods. Furthermore, GenoGraph presents a versatile framework to identify key genetic variants that can be generalized to other traits and phenotypes, offering a powerful and adaptable solution for identifying genetic variants associated with complex phenotypes.

## DESIGN

The overall framework of GenoGraph is illustrated in Figure 1. We utilized PLINK^23^ software for genotype quality checking and standard normalization techniques as pre-preprocessing steps and generated a genotype matrix 𝑋 ∈ ℝ^n×d^, with 𝑑 number of variants and 𝑛 individuals (Figure S1). Subsequently, we constructed the graph 𝒢, with nodes representing the individuals and the edges representing the relations between the individuals (nodes) with genetic similarity, using the weighted Hamming distance.^24^ A detailed description of the data pre-processing and graph representation can be found in the Methods details section.

**Figure 1.**
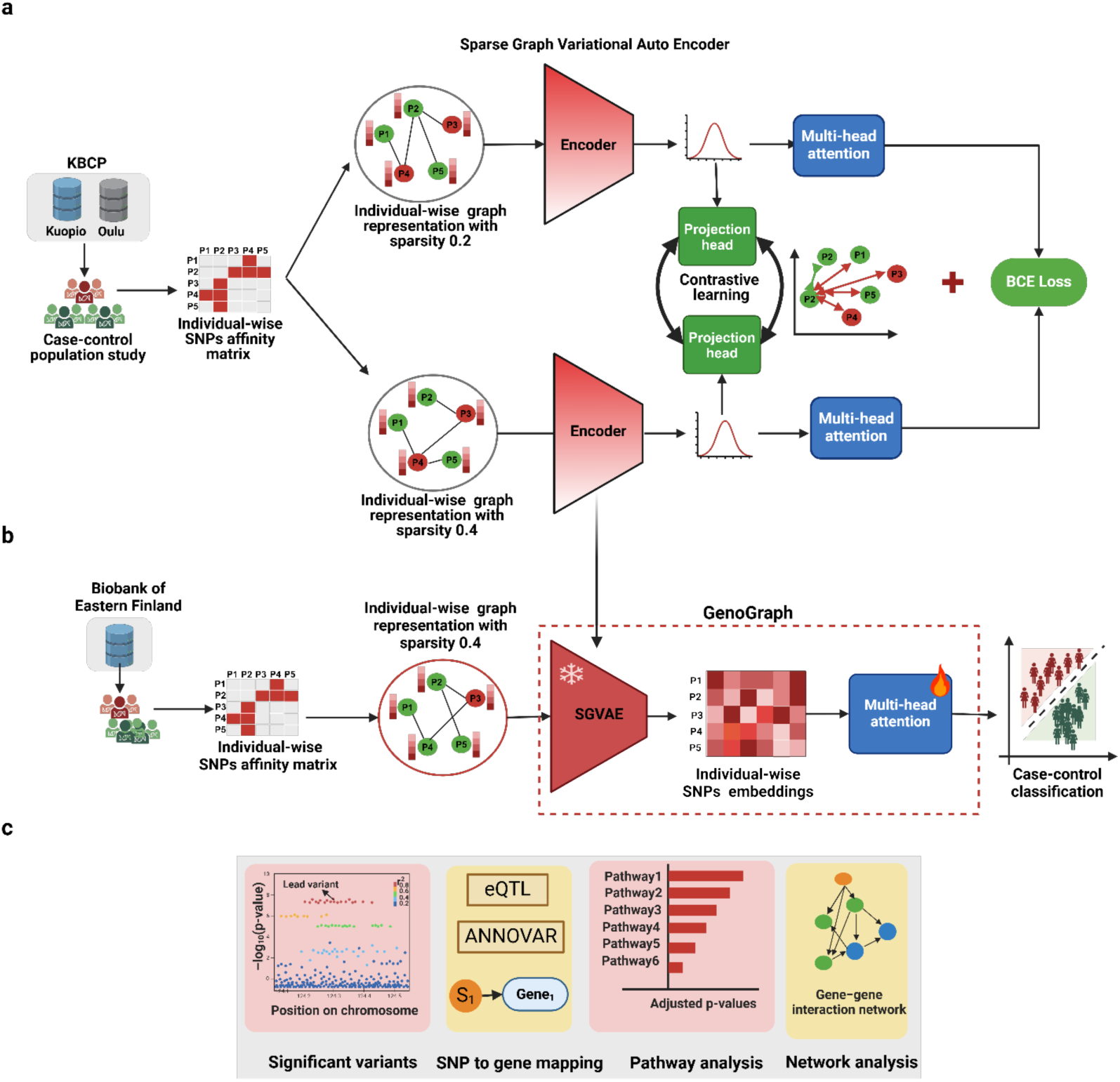
An interpretable graph-based contrastive learning approach for variant representation learning in a BC risk-prediction study. **a**, architecture of the proposed Sparse Graph Variational Autoencoder network combined with a supervised contrastive learning approach (GenoGraph) for variant representation learning. The framework begins with two augmented versions of the input graph 𝒢 with individuals as nodes and their genetic similarity as edges, processed through an encoder block comprising three graph-convolution–based message-passing layers. The encoder leverages the re-parameterization trick from variational autoencoders (VAEs) to generate latent space vectors. These vectors are passed through a projection head to generate variant embeddings and are supervised using a multi-head attention classifier within the contrastive learning optimization technique. **b**, Fine-tuned GenoGraph framework, which uses the pre-trained weights to generate variant embeddings from the input graph and employs a multi-head attention-based classifier to perform graph node classification tasks. **c**, Interpretation: biological relevance pipelines to compute variant importance scores to identify the influential variants, functionally annotate and gene map the identified variants, and apply gene-set enrichment analysis to identify the pathways related to BC risk and interaction networks. Finally, gene–gene interaction networks are inferred to interpret the biological roles of the identified genetic variants.

In the pre-training phase (Figure 1a), we trained the proposed SVGAE framework aimed at learning variant embeddings by incorporating a supervised contrastive learning technique.^20,21^ We employed edge perturbation with sparsity thresholds 0.2 and 0.4 to generate two augmented version of graph 𝒢, referred to as 𝒢*_1_* and 𝒢*_2_*.^25^ The core of the SGVAE encoder consists of three graph convolutional layers, to propagate information between the neighboring nodes while updating each node’s embeddings based on its local neighborhood.^26,27,28^ Layer normalization^29^ and dropout^30^ with 𝑝 = 0.3 were strategically applied between the convolution layers to stabilize training and prevent overfitting. The re-parametrized latent representation 𝑍 ∈ ℝ^n×5^^12^ captures each individual (node) variant representation by considering the underlying graph structure. The latent space embeddings 𝑍 ∈ ℝ^n×5^^12^ were further projected through a two-layered fully connected network (FCN), (projection head) with 256 and 128 dimensions, respectively, represented as 𝑍*_proj_* ∈ ℝ^n×1^^28^.

### Fine-tuning GenoGraph for downstream analysis

We fine-tuned the GenoGraph framework and subsequently used the pre-trained SVGAE encoder for node classification in a case-control prediction study (Figure 1b). To interpret model predictions, we applied GNNExplainer as a post-hoc interpretability technique, enabling the identification of the most influential genetic variants.^22^ The top-ranked variants were further investigated to assess their biological relevance and potential contribution to BC susceptibility (Figure 1c). A detailed description of the pre-trained and fine-tuned loss functions are provided in the Method details section.

## RESULTS

We used three genetic variant datasets, Kuopio Breast Cancer Project (KBCP) (𝑛 = 696, cases = 445, controls = 251), Oulu Breast Cancer Study (OBCS) (𝑛 = 923, cases = 508, controls = 415), and the Biobank of Eastern Finland (BCAnalyze) (𝑛 = 1726, cases = 526, controls = 1200). These datasets were obtained with their respective approvals from Kuopio and Oulu University Hospital and the appropriate ethics committees, and all the participants provided written informed consent prior to this study. To ensure data privacy and participants’ confidentiality, only de-identified data were used. All experiments involving these datasets adhered to the relevant ethical guidelines outlined in the Declaration of Helsinki.

### A pretrained GenoGraph framework demonstrates superior performance over graph-based message-passing operations

To evaluate the efficacy of a pretrained GenoGraph framework, we compared its performance against various graph-based message-passing operations, including GCNConv^26^, GATConv^27^, and SAGEConv^28^. The graph models were pretrained using the combined training datasets from the KBCP and OBCS datasets, adhering to the data-splitting protocol detailed in the Method details section. Subsequently, their performance was evaluated on independent test sets from the KBCP, OBCS, and BCAnalyze datasets to assess efficacy in a BC case–control classification. The results, depicted in Figure 2, provide an in-depth comparison across three key performance metrics: accuracy, AUC-ROC, and AUC-PRC. GenoGraph, which integrates GCNConv with layer normalization^29^ and dropout^30^, consistently outperformed other graph-based convolution methods across all metrics. This enhanced performance underscores the importance of incorporating normalization and regularization techniques for representation learning in HDLS scenarios.

**Figure 2.**
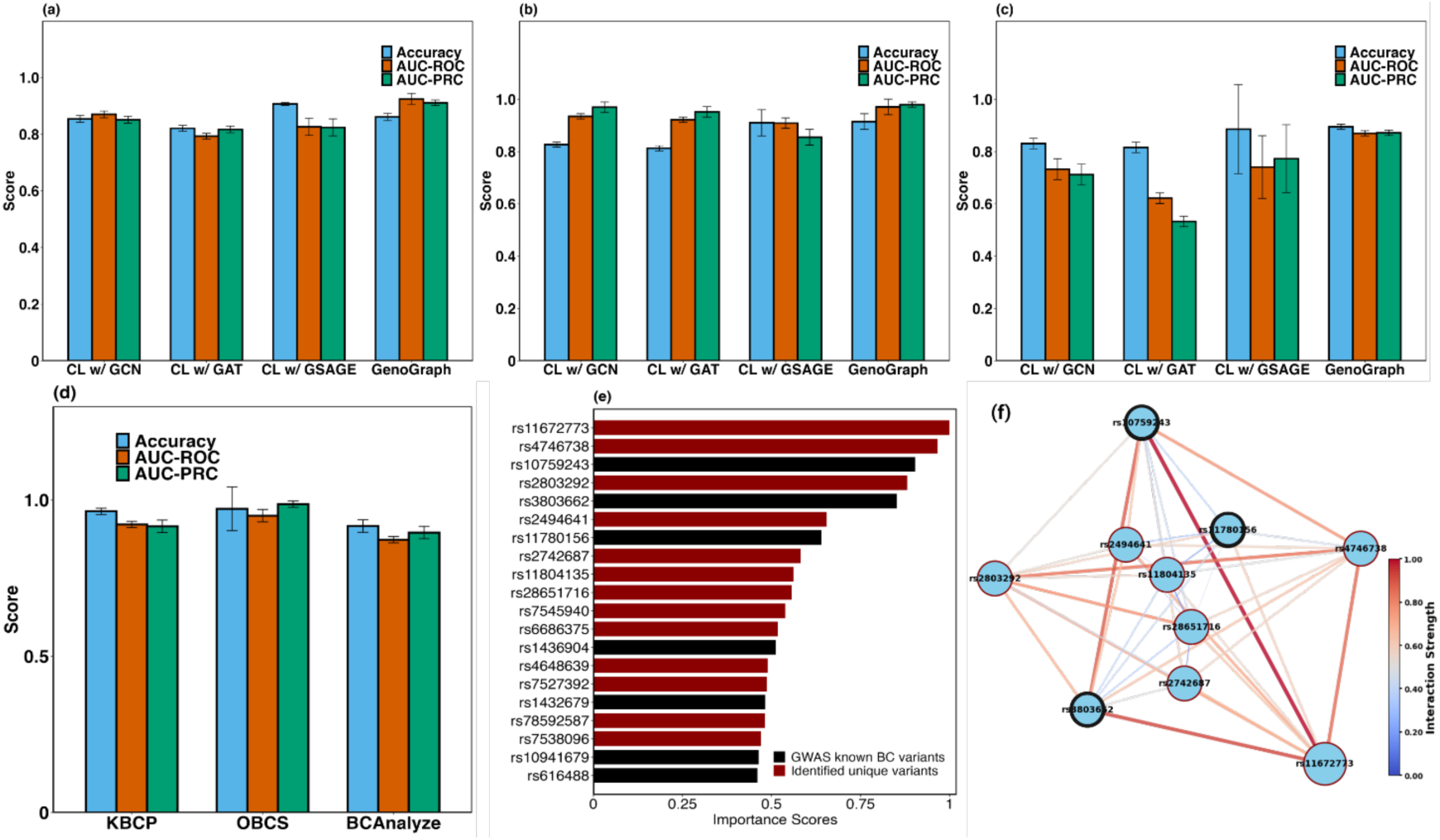
Predictive performance of the pretrained and fine-tuned GenoGraph framework. **a, b, c**, Breast cancer case–control classification performance of various graph-based message-passing operations pre-trained with a supervised contrastive learning approach on the test sets of KBCP, OBCS, and BCAnalyze, respectively. **d**, Evaluation of the fine-tuned GenoGraph framework for case–control classification tasks. **e**, Contribution of the top 20 variants identified using the fine-tuned GenoGraph framework, where black bars indicate variants previously associated with BC risk in the literature. **f**, Visualization of the variant–variant interaction network, where interaction strengths were computed using the self-attention module within the GenoGraph framework. Nodes outlined in thick black denote variants known to be associated with BC risk. The abbreviations in the figure are CL: contrastive learning; w/: with; GCN: GCNConv; GAT: GATConv; SAGE: SAGEConv; GenoGraph: CL w/ GCNConv followed by Layer normalization and dropout.

On the KBCP test set (Figure 2a), GenoGraph demonstrated substantial improvements over other graph convolution models across all metrics. Specifically, GenoGraph achieved an AUC-ROC of 0.95, a relative improvement of 2.7% compared with SAGEConv. In terms of AUC-PRC, GenoGraph exhibits a 2.15% relative improvement over SAGEConv. Additionally, the case–control classification accuracy of GenoGraph was 3.3% higher compared with SAGEConv. These improvements highlight the framework’s capacity to effectively capture predictive genetic variants, leading to enhanced BC classification performance. Evaluating on the OBCS test dataset further validated these findings (Figure 2b), GenoGraph achieved an AUC-ROC of 0.96, exceeding 0.94 and 0.93 for SAGEConv and GATConv, respectively. This corresponds to a relative improvement of 2.13% over SAGEConv. On the independent BCAnalyze dataset (Figure 2c), GenoGraph demonstrated modest yet consistent improvements over other models. Specifically, GenoGraph achieved an AUC-ROC of 0.92, compared with 0.91 for SAGEConv, with a relative improvement of 1.1%. In terms of AUC-PRC, GenoGraph outperformed SAGEConv by 1.76%. Similarly, GenoGraph’s accuracy was 2.27% higher than SAGEConv’s. Although the gains were less pronounced compared with those seen with the KBCP and OBCS datasets, GenoGraph’s consistent outperformance across all metrics suggests its capability for generalization, even under more challenging conditions.

### Enhanced predictive accuracy for breast cancer case–control classification through fine-tuned GenoGraph

Fine-tuning of the pre-trained GenoGraph model, specifically leveraging the SGVAE encoder to generate variant embeddings, demonstrated enhanced improvements in BC case–control classification. These variant embeddings were subsequently used within a self-attention block, allowing us to evaluate the downstream utility of pre-trained variant representations.

The fine-tuned GenoGraph model exhibited substantial enhancements in classification accuracy across three independent test datasets—KBCP, OBCS, and BCAnalyze (Figure 2d). Specifically, relative improvements in predictive accuracy of 11.69%, 6.23%, and 2.48% were achieved using the test sets from KBCP, OBCS, and BCAnalyze, respectively, when compared with the pre-trained GenoGraph model. Similarly, improvements in the AUC-PRC were observed, with relative improvements of 0.54%, 0.71%, and 2.75% for KBCP, OBCS, and BCAnalyze, respectively. Although a negative relative improvement of 2.62% in AUC-ROC was observed for the fine-tuned model with the OBCS dataset, the overall performance indicates a substantial improvement, particularly in capturing meaningful variant embeddings for BC risk assessment.

The robustness of the fine-tuned model was further highlighted by its performance on the BCAnalyze dataset, which is characterized by substantial variability and complexity. Despite these challenges, GenoGraph achieved an accuracy of 0.917 ± 0.02, an AUC-ROC of 0.873 ± 0.01, and an AUC-PRC of 0.896 ± 0.02. Notably, these results represent an average improvement of 6.83% in accuracy and 1.33% in AUC-PRC compared with models without pre-training, underlining the capability of the pretrained GenoGraph framework to generalize effectively even under complex conditions.

#### Identification of interpretable and interacting genetic variants by GNNExplainer

We used the GNNExplainer to interpret and compute the importance scores of genetic variants within the fine-tuned GenoGraph framework on the BCAnalyze test set. We determined the median importance scores across all individuals to calculate global feature importance scores, which facilitated the identification of the most influential variants (Supplementary Document S2, Table 1). The top 20 influential variants identified by the GenoGraph framework, ranked by their global importance scores, highlighted the critical subset of SNPs that potentially contribute for BC risk (Figure 2e). Notably, seven of these top variants, depicted by black bars, have previously been associated with BC risk in various GWASs.^10^

Additionally, to explore the intricate complex relationships among genetic variants, we used a self-attention mechanism within the GenoGraph framework to compute interaction strengths among the top identified variants (Supplementary Document S2, Table 2). The network visualization of variant–variant interactions focused on the top key significant variant, rs11672773, demonstrating its strong interaction with other BC risk variants (Figure 2f). In the network, the nodes within the black contour are previously identified BC risk variants.^10^ The SNP rs11672773 (*ZNF8*) exhibits a robust interaction with rs10759243 (*KLF4*) and rs3803662 (*TOX3*), with interaction strengths of 0.99 and 0.92, respectively. *ZNF8* has been identified as a key player in BC progression, particularly in modulating metastasis through the transforming growth factor-β (*TGF-β*) pathway by its interaction with the gene *SMAD3*, enhancing the expression of genes linked to BC lung metastasis.^31^ Krüppel-like factor 4 (*KLF4*) is a zinc finger-containing transcription factor essential for regulating cell growth, proliferation, and differentiation, and its strong interaction with *ZNF8* underscores its critical role in cancer biology.^32^ *TOX3* influences the hormonal and cellular pathways involved in BC, although the exact mechanisms are unclear.^33^ *TOX3* interaction with both *ZNF8* and *KLF4* in the network suggests a complex interplay that might influence key pathways in BC progression and manifestation. The elucidation of variant–variant interaction networks underscore the efficacy of GenoGraph to identify and visualize important genetic interactions that could be vital for further research and therapeutic development.

#### GenoGraph demonstrates superior performance compared with existing machine- and deep learning–based GWAS approaches

We systematically evaluated subsets of SNPs comprising 100, 500, 1000, 2000, 2500, and 5000 top-contributing variants identified by the GenoGraph framework to determine the optimal subset for further biological relevance analysis in the context of BC risk pathways. Logistic regression was employed as the case-control predictive classifier, and the performance of these SNP subsets was assessed across the test sets of the KBCP, OBCS, and BCAnalyze datasets.

An optimal subset of 2500 SNPs was consistently identified for the KBCP and BCAnalyze datasets, yielding significant performance improvements. Specifically, its use achieved relative improvements of 8.60% and 10.68% in accuracy over the average accuracy of all evaluated subsets for KBCP (Figure 3a) and BCAnalyze (Figure 3c), respectively. In contrast, the OBCS dataset exhibited superior performance with a subset of 2000 SNPs, achieving a 10.68% relative improvement in accuracy compared to the average performance across all subsets (Figure 3b). Beyond these optimal set sizes, further increases in the SNP subset led to a decline in performance metrics, demonstrating a clear threshold in predictive accuracy and model efficiency. This finding underscores the importance of determining an optimal SNP subset size that balances predictive performance with computational efficiency; this balance is crucial in the practical application of genetic data analyses in clinical settings. We used these optimal subsets for further analysis, focusing on elucidating the biological relevance of these SNPs in relation to BC risk pathways.

**Figure 3.**
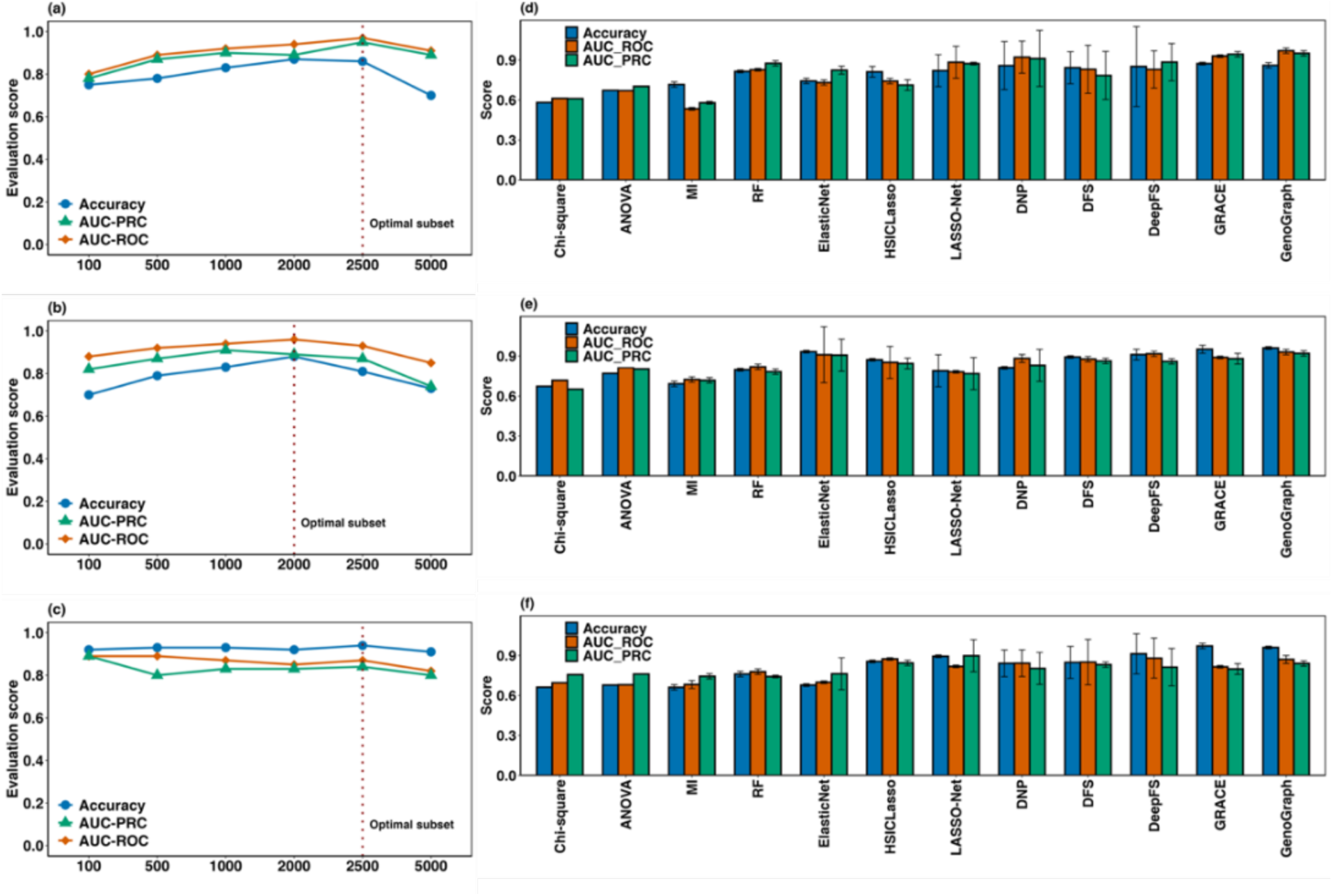
Comprehensive evaluation of an optimal SNP subset using the GenoGraph framework and existing approaches. **a, b, c**, Evaluation of the logistic regression classifier to find the optimal subset of SNPs, using GenoGraph framework–identified variants, on the test sets of KBCP, OBCS, and BCAnalyze, respectively. **d, e, f**, Comparative analysis of the optimal subset of variants identified by a range of statistical, ML, DL, and graph-based deep-learning approaches relative to the GenoGraph framework, on the test sets of KBCP, OBCS and BCAnalyze, respectively.

Additionally, we evaluated the predictive capabilities of various HDLS methods derived from statistical, ML, and DL approaches, including chi-square, ANOVA, mutual information (MI)^34^, RF^35^, ElasticNet^36^, HSIC Lasso^37^, LASSONet^38^, Deep Neural Pursuit (DNP)^39^, Deep Feature Selection (DFS)^40^, Deep Feature Screening (DeepFS)^41^, and GRCAES^42^ as baseline methods. These methods were employed to ascertain the performance of the optimal subset of SNPs identified by each approach, in comparison with GenoGraph.

We employed optuna to find the optimal hyper-parameters for statistical and baseline ML approaches, which were subsequently used to train logistic regression classifiers. For DL methods, we adopted the default configurations as specified in their original publications. We selected the top 2,500 contributing SNPs on the basis of computed feature importance scores, trained, and validated a logistic regression classifier on these SNPs. GenoGraph exhibited a notable enhancement in performance across multiple test sets when evaluated against the graph-based FS method GRACE. Specifically, GenoGraph achieved a relative improvement in terms of AUC-ROC of 4.22%, 7.15%, and 6.32% for the KBCP (Figure 3d), OBCS (Figure 3e), and BCAnalyze (Figure 3f) datasets, respectively. Despite these improvements in AUC-ROC, GRACE demonstrated superior performance in terms of accuracy, with particularly notable relative improvements of 1.51% and 1.14% on the KBCP and BCAnalyze datasets, respectively. This differential in performance metrics between the two methods underscores the importance of selecting appropriate FS strategies on the basis of specific needs, suggesting a potential complementarity of GenoGraph and GRACE for enhancing predictive accuracy and model robustness in HDLS data.

The comparative analysis of various FS methods using HDLS data is illustrated in Figure S2, with each method distinguished by unique shapes within its category and differentiated by color. Methods named near the top right of the figure exhibited optimal stability and performance, while those toward the bottom left yielded suboptimal characteristics. The statistical methods, including chi-square, ANOVA, and MI, though characterized by high stability, yielded lower performance compared with other FS categories. These methods are highlighted in the black dotted box. Conversely, ML methods such as RF and Elastic Net exhibited both low stability and performance, suggesting limitations in handling HDLS data effectively. DL-based methods, including HSIC Lasso, LASSONet, DNP, DFS, and DeepFS, generally offered superior performance. However, they are challenged by stability issues, except for DeepFS, which demonstrated higher stability in line with slightly lower performance relative to other DL methods. This balance between performance and stability is encapsulated within the red dotted box in Figure S2. Graph-based approaches, particularly GRACE and GenoGraph, stand out by achieving both high performance and stability. Despite this, there exists a noticeable trade-off between stability and performance across all methods, indicating that none of the FS approaches can guarantee concurrent high stability and high performance. While DL-based methods consistently manage the trade-off between stability and performance, as highlighted by the red dotted box, GenoGraph emerges as a method with exceptional stability but relatively lower performance when compared with DeepFS. This insight underscores the complex dynamics between stability and performance in FS methods, especially in the context of HDLS datasets.

#### GenoGraph identified 370 breast cancer–associated variants in the Finnish population

We employed logistic regression to the GenoGraph-identified optimal subset of SNPs (2,500) to identify BC-associated genome-wide significant variants at 𝑝 < 5 × 10^,+^, using Bonferroni correction to account for multiple hypothesis testing (Supplementary Document S2, Table 3). Of 2,500 SNPs, 370 were found to be significant, among which 85 SNPs are found to be associated with BC risk in the GWAS catalog (Figure S3). Out of the 2,500 variants initially identified, 400 variants were missing rsID information, resulting in the annotation and gene mapping of 2,100 variants using SNPxplorer.^43^ The genetic variants identified by GenoGraph reside predominantly within noncoding regions, with the majority being intergenic (26.86%) and intronic (25.48%) (Figure 4a) (Supplementary Document S2, Table 4). Our findings suggest that these variants may influence gene regulation, potentially through long-range regulatory elements or splicing mechanisms. Variants within regulatory regions (23.38%) could affect gene expression by altering enhancer or promoter activities, which is critical in disease contexts like BC. A smaller fraction of variants were identified in coding regions, including missense (1.9%) and synonymous variants (0.52%), which may alter protein function or gene expression stability. These results emphasize the significant role of both coding and noncoding variants in influencing gene regulation and expression.

**Figure 4.**
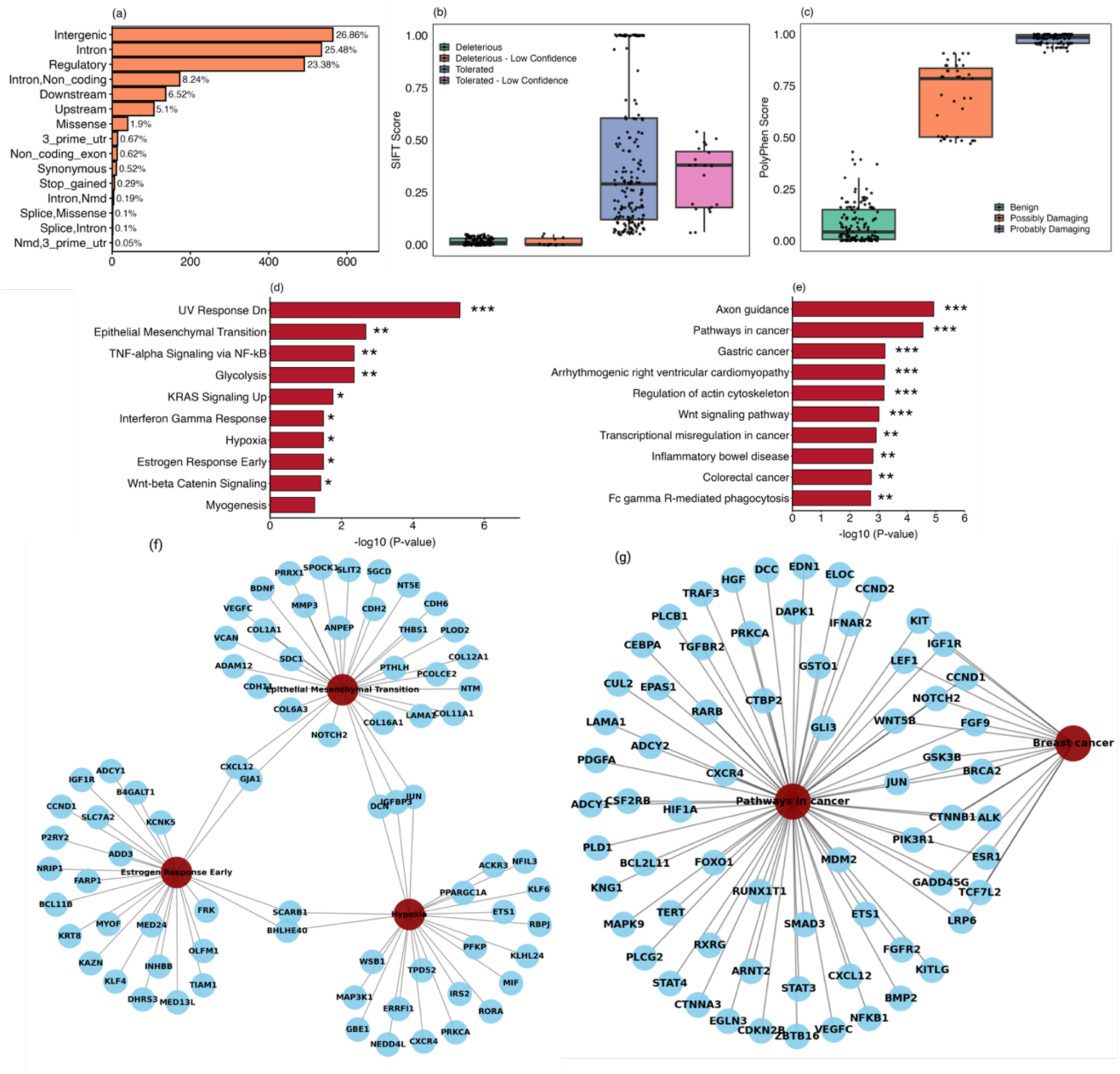
Functional annotation of variants, enrichment analysis, and tissue specific GTEx analysis for variants identified by the GenoGraph framework. **a**, Distributions of SNP positions and consequences across different genomic regions, highlighting the types of variants identified, with a notable presence of intergenic, intronic, and regulatory region variants. **b**, Distribution of SIFT scores, categorizing variants on the basis of their predicted impact on protein function. **c**, Distribution of PolyPhen scores, showing the predicted impacts of variants on protein stability as "benign", "possibly damaging", or "probably damaging". **d, e**, Enriched pathways from the MSigDB Hallmark 2020 and KEGG gene sets ranked by −log10 adjusted p-values, respectively. **f**, Network illustration of three enriched pathways—EMT, Estrogen Response Early, and Hypoxia—illustrating their associated genes in MSigDB. **g**, Network representation of the pathways in cancer and their associated genes, with a specific emphasis on breast cancer-related genes in the KEGG gene set database.

Sorting Intolerant From Tolerant (SIFT)^44^ scores predict whether amino acid substitutions affect protein function, with lower scores indicating a higher probability of deleterious effects (Supplementary Document S2, Table 5). Notably, 41.03% of the variants are classified as deleterious, with scores close to zero, suggesting a significant potential for disrupting protein function. A smaller group (3.42%) is also classified as deleterious but with low confidence, indicating uncertainty likely due to limited supporting data. In contrast, 49.86% of the variants are classified as tolerated, implying a lower likelihood of affecting protein function, while 5.7% are tolerated with low confidence (Figure 4b). These findings emphasize the need to consider both high-confidence predictions and potential uncertainties when interpreting the functional relevance of genetic variants.

The PolyPhen^45^ score evaluates the impact of missense mutations on protein stability and function based on evolutionary conservation, structural context, and biochemical properties (Supplementary Document S2, Table 6). In our analysis, 47.33% of the variants are classified as benign, indicating a minimal likelihood of affecting protein function, as evidenced by low PolyPhen scores. Conversely, 38.79% are deemed probably damaging, with high scores suggesting a significant potential to disrupt protein function, which may be crucial for understanding disease mechanisms. Additionally, 13.88% are categorized as possibly damaging, with moderate scores that indicate a potential functional impact under certain conditions (Figure 4c). This distribution highlights the varied effects of the variants on protein functionality and the importance of further study of their biological significance and potential roles in disease phenotypes.

### Pathway enrichment analysis reveals distinct biological process associated with breast cancer

We conducted a gene set enrichment analysis (GSEA)^46^ for 2,100 genes mapped to the GenoGraph- identified optimal subset of SNPs. Our findings reveal distinct pathways associated with BC risk (Supplementary Document S2, Table 7). Figure 4d shows the top 10 enriched pathways from analysis of the MSigDB gene set. Notably, UV Response Dn exhibited the highest level of significance (adjusted 𝑝 = 1.63 × 10⁻⁸), suggesting a notable cellular response to UV-induced stress, potentially linked to genomic instability, which is a key driver of cancer progression, including in BC. UV radiation induces DNA damage that can lead to cancer progression through various pathways, including the activation of autophagy signaling and the *p53* tumor suppressor pathway. Autophagy plays a critical role in cellular homeostasis and can influence cancer progression by regulating genomic integrity under stress conditions, such as those induced by UV exposure^47^. The *p53* pathway, which is crucial for DNA repair and cell cycle regulation in response to UV damage, also highlights the potential for UV exposure to contribute to genomic instability and cancer development.^48^ Enrichment of the pathways Epithelial Mesenchymal Transition (EMT) (adjusted 𝑝 = 8.88 × 10⁻⁴) and Estrogen Response Early (adjusted 𝑝 = 4.14 × 10⁻²) indicates their potential involvement in BC progression, with EMT’s enhanced cellular motility and plasticity being particularly critical for cancer metastasis.^31^

Figure 4f is the network representation of the three enriched pathways—EMT, Estrogen Response Early, and Hypoxia—along with their associated genes. Within the Estrogen Response Early pathway, genes such as *IGF1R* and *CCND1* are involved in cell proliferation and hormone signaling, which are pivotal in the development of hormone receptor–positive BC. In the EMT pathway, genes like *NOTCH2* and *COL11A1* are involved in cellular differentiation and extracellular matrix remodeling, which play essential roles in metastasis. Furthermore, the Hypoxia pathway highlights genes such as *HIF1A* and *VEGFC*, which regulate angiogenesis and cellular adaptation to low oxygen, contributing to tumor aggressiveness and resistance to therapies.

Figure 4e and 4g illustrate findings from the KEGG database pathways associated with the identified variants. Figure 4e presents the top 10 enriched pathways, with Axon Guidance being the most significant, followed by Pathways in Cancer, Transcriptional Misregulation in Cancer, and the Wnt signaling pathway. These pathways are indicative of core molecular mechanisms in oncogenesis, including cellular communication, transcriptional regulation, and signaling processes that drive cancer development. Figure 4g presents a network of the pathways in cancer and their associated genes, with an emphasis on genes directly linked to BC risk. Key genes such as *BRCA2, ESR1*, and *PIK3R1* are featured, underscoring their importance in BC risk and pathogenesis.^49,50,51^ *BRCA2* is a well-known tumor suppressor gene, mutations of which significantly elevate BC risk. *ESR1* plays a crucial role in estrogen-mediated growth of hormone receptor-positive BC, while *PIK3R1* is implicated in oncogenic signaling through its role in cell growth and survival.

Although the pathways in cancer finding was statistically significant, the specific BC pathway itself did not reach statistical significance in this analysis, with an adjusted 𝑝 = 0.17. However, the presence of well-established BC-associated genes within significant pathways suggests that the GenoGraph framework successfully identified variants involved in crucial regulatory mechanisms of BC. This highlights the relevance of the identified variants in cancer biology, even though direct statistical significance for the BC pathway was not attained, reflecting a potential limitation in statistical power or pathway representation rather than biological irrelevance. Overall, these findings underscore the ability of GenoGraph to identify genetic variants linked to key oncogenic processes, thereby providing insights into the molecular underpinnings of BC, albeit without conclusive direct evidence for all pathways at stringent significance thresholds.

#### Analyzing tissue-specific differential gene expression in breast cancer and performing enrichment analysis of associated pathways

The analysis of tissue-specific differentially expressed genes (DEGs) across 30 different tissue types revealed significant insights into the transcriptional landscape of BC (Figure 5a). DEGs were categorized as upregulated (DEG.up, shown in green) (Supplementary Document S2, Table 8), downregulated (DEG.down, shown in orange) (Supplementary Document S2, Table 9), and nondirectional significant changes (DEG.twoside, shown in blue). For breast tissue, 171 of the 1,978 upregulated genes demonstrated significant associations (Bonferroni-adjusted 𝑝 = 5.58 × 10^,%-^), indicating potential roles in BC pathogenesis through enhanced gene activity. Of the 1,007 downregulated genes, 77 were significantly associated with breast tissue (adjusted 𝑝 = 7.57 × 10^,$^), implying involvement in loss of tumor-suppressive functions. Moreover, 248 of the 2,985 genes with two-sided differential expression were significantly linked to breast tissue (adjusted 𝑝 = 1.48 × 10^,&%^), underscoring a complex regulatory network characterized by both gene activation and suppression (Supplementary Document S2, Table 10).

**Figure 5.**
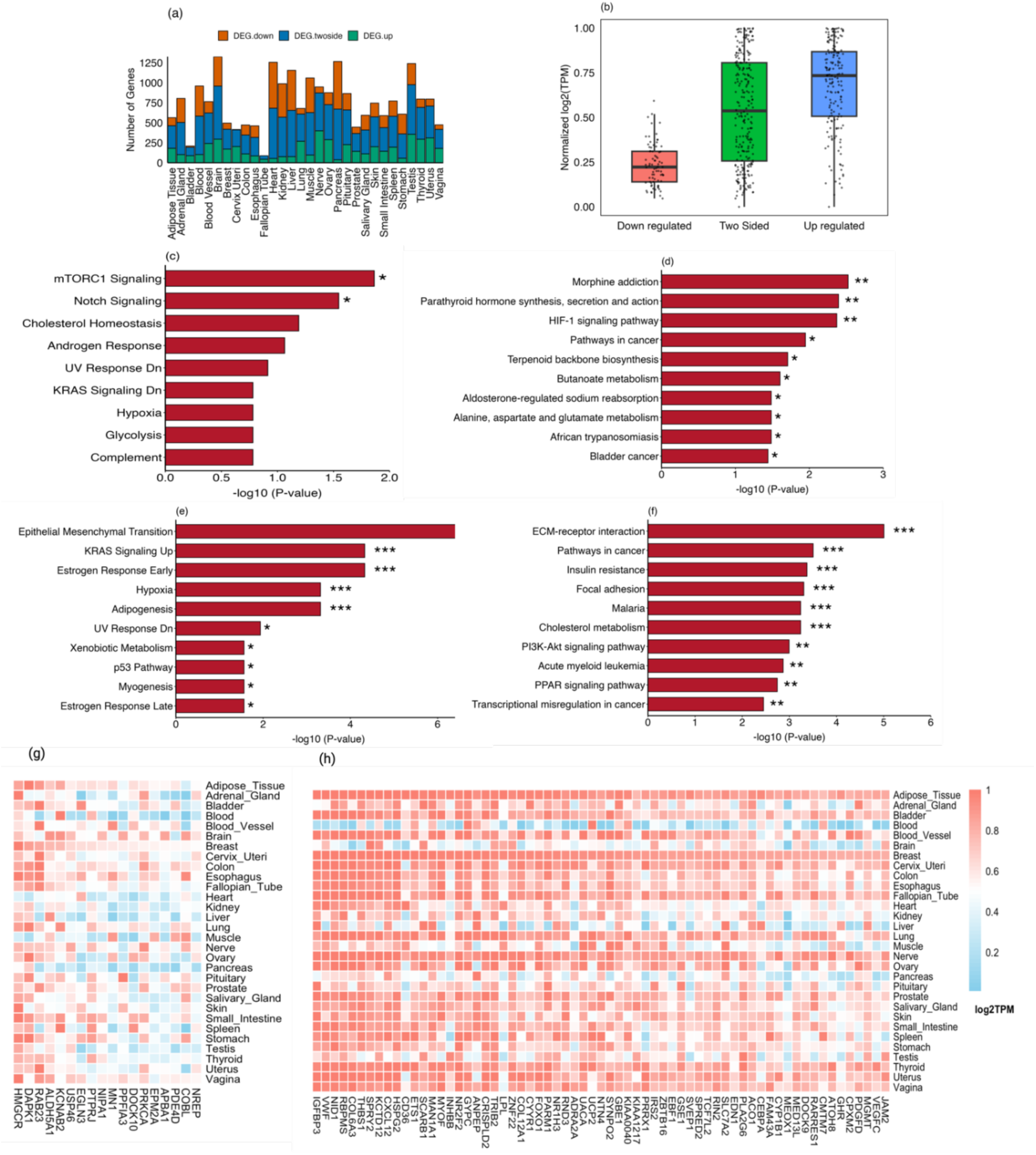
Pathway enrichment analysis using GSEA reveals significant biological processes associated with differentially expressed genes in breast tissue. **a**, Tissue-specific distribution of differentially expressed genes (DEGs) across various tissue types, showing the number of DEGs that are upregulated, downregulated, or two-sided in each tissue. **b**, Gene expression in breast tissue, represented as a box plot of normalized expression (log2TPM) values. The plot categorizes gene expression as "downregulated", "two-sided", or "upregulated", revealing distinct expression patterns in breast tissue. **c, d**, Top enriched pathways from the MSigDB and KEGG gene sets, respectively, ranked by −log10 p-values for the downregulated DEGs in breast tissue. **e, f**, Top enriched pathways from the MSigDB and KEGG gene sets, respectively, ranked by −log10 p-values for the upregulated DEGs in breast tissue. **g, h**, Heatmap of gene expression across multiple tissue types, representing the normalized TPM scores of key downregulated and upregulated DEGs, respectively.

Among the 171 upregulated and 77 downregulated differentially expressed genes (DEGs), 38% and 23.38%, respectively, exhibited elevated transcript activity, with transcripts per million (TPM) values exceeding 0.8 (Supplementary Document S2, Table 11). These findings suggest significant overexpression and potential roles in driving breast cancer progression, further highlighting their critical regulatory functions (Figure 5b). We further explored the biological roles of the 23.38% of downregulated DEGs and 38% of upregulated DEGs with elevated TPM scores above 0.6 and 0.8, respectively. A GSEA using the MSigDB and KEGG pathway databases revealed significant enrichment of pathways related to BC progression. For downregulated DEGs, the most prominent pathways were *mTORC*1 and Notch signaling (Figure 5c), with adjusted 𝑝 −values of 0.123 and 0.127, respectively. These pathways have well-documented involvement in tumor growth, metabolism, and differentiation in BC. The *mTORC1* pathway, a regulator of cell metabolism and proliferation, showed enrichment with the genes *EGLN3* and *HMGCR. EGLN3*, encoding a regulator of the hypoxic response, and *HMGCR*, encoding a key enzyme in cholesterol biosynthesis, were both downregulated. This may indicate tumor cells’ reduced ability to manage hypoxic stress and cholesterol synthesis, which may limit their proliferation.^52,53,54,55,56^

The association of 28% of the downregulated genes with breast and other tissues is presented in Figure5g, highlighting the tissue-specific relevance of genes such as *HMGCR, DAPK1*, and *RAB23*. These genes exhibited high transcript activity (TPM > 0.8) and are significantly associated with BC. *HMGCR* is involved in cholesterol biosynthesis, supporting tumor proliferation, while *DAPK1* acts as a tumor suppressor. *RAB23* is implicated in cancer cell invasion and metastasis. Although *ALDH5A1* and *KCNAB2* are less directly linked to BC, their roles in cancer stem cell regulation and apoptosis suggest broader relevance. Together, these findings underscore the complex regulatory mechanisms of downregulated genes in BC pathophysiology.

The EMT pathway from MSigDB was the most enriched for the upregulated DEGs, with an adjusted 𝑝 −value < 0.000008, emphasizing its role in BC progression of promoting cellular plasticity, invasion, and metastasis (Figure 5e). The Estrogen Response Early and *KRAS* signaling pathways were also significantly enriched (𝑝 < 0.0005), indicating the involvement of early estrogen signaling in hormone receptor-positive BCs and oncogenic *KRAS* signaling in promoting tumor proliferation and survival. Additional pathways included Hypoxia and Adipogenesis, both with adjusted 𝑝 < 0.003, highlighting the importance of these mechanisms in the tumor microenvironment and altered lipid metabolism, respectively.

The pathways ECM-receptor interaction, Pathways in Cancer, Insulin Resistance, Focal Adhesion, and Cholesterol Metabolism were significantly enriched in the upregulated DEGs, with adjusted 𝑝 −values of 0.011 (Figure5f). The association of these 65 genes with BC is also depicted in the heatmap, which includes 30 other tissue types (Figure 5h). Specifically, six genes—*CXCL12, CD36, VEGFC, THBS1, CYIB1,* and *TCF7L2*—demonstrated strong evidence for association with BC. *CXCL12* plays a crucial role in tumor progression, metastasis, and immune response, being implicated in breast cancer cell migration and invasion.^57^ *CD36*, which is involved in lipid metabolism, may support the cancer stem cell population, contributing to chemoresistance and metastasis.^58,59,60,61^ *VEGFC*, a gene associated with angiogenesis, plays a critical role in tumor growth and metastasis.^62,63^ *THBS1* is involved in cell–cell and cell–matrix interactions, with both pro- and anti-tumorigenic effects, depending on the context, and has been linked to BC development.^64,65,66^ *CYIB1*, involved in estrogen metabolism, is associated with hormone-dependent cancers, including BC.^67,68,69^ Lastly, *TCF7L2*, part of the Wnt signaling pathway, is associated with BC risk, underscoring its importance in tumorigenesis.^70,71,72^ Our findings illustrate the intricate tissue-specific mechanisms of gene regulation in BC and highlight multiple potential pathways and gene targets for future therapeutic strategies. The Notch signaling pathway, frequently dysregulated in BC, involves cell differentiation and proliferation. The downregulation of *PRKCA* in our analysis suggests a reduction in this pathway’s activity, potentially leading to diminished tumorigenic processes.^73^

KEGG pathway analysis revealed moderate enrichment in the morphine addiction and *HIF-1* signaling pathways (Figure 5d). *PDE4D* and *PRKCA*, both implicated in the morphine addiction pathway, are involved in *cAMP* signaling, and their downregulation could affect tumor cell behavior by disrupting cellular signaling. Crosstalk between neurotransmitter signaling and cancer biology is increasingly recognized as a contributor to tumor progression.^74,75^ The *HIF-1* signaling pathway, critical for the tumor response to hypoxia, showed downregulation of *EGLN3* and *PRKCA*, which may impair the hypoxic response and reduce tumor survival in low-oxygen environments.^76^

## DISCUSSION

In this work we developed GenoGraph, a graph-based contrastive learning framework that effectively identifies BC risk variants and provides biologically relevant insights into their roles. By leveraging a supervised graph-representation learning approach, GenoGraph addresses the inherent challenges of high-dimensional, low-sample-size genetic data, surpassing the performance of conventional GWAS, ML, and DL methodologies. Compared with traditional ML and DL approaches, GenoGraph demonstrates several advantages. GenoGraph excels by modeling intricate relationships between genetic variants and identified 2,500 crucial variants, 370 of which are significantly associated with BC risk.^10^

Recent GWAS studies, including large meta-analyses and cross-ancestry evaluations, have identified numerous novel BC risk loci, expanding our understanding of the genetic predisposition to BC.^77^ However, these traditional approaches have inherent challenges in incorporating genetic interactions. The unique contribution of GenoGraph lies in its ability to not only identify significant variants but also interpret the biological relevance of their interactions, which is often challenging for conventional GWAS. Such interpretability is crucial, as exemplified by pathways like EMT, a process associated with cancer metastasis that has been highlighted in previous GWASs for its role in BC progression.

One significant advantage of GenoGraph compared with ML and DL methods is its resilience to overfitting, a major issue when dealing with HDLS genetic datasets. While DL approaches like DeepGWAS integrate regulatory information, they still suffer from stability issues under HDLS conditions. GenoGraph’s use of graph representation learning, and multi-head attention mechanisms tackles these challenges directly, providing enhanced predictive accuracy and stability in HDLS contexts. It also stands out for its consistently superior performance across multiple datasets, which suggests strong potential for generalizability—a major goal in precision medicine research.

The consistent enhancement across diverse datasets underscores the utility of pre-trained variant embeddings for downstream BC classification tasks. The SGVAE encoder, by leveraging pre-training, effectively captured variant-specific information that retained discriminative power, facilitating improved predictive performance. Across all three datasets, GenoGraph consistently outperformed GCNConv, GATConv, and SAGEConv in accuracy, AUC-ROC, and AUC-PRC. The average relative improvement observed was 2–3% in AUC-ROC and accuracy, and 1–2% in AUC-PRC. These enhancements are particularly notable given the intrinsic challenges posed by high-dimensional genomic data. Layer normalization and dropout played a vital role in stabilizing the training process, reducing susceptibility to overfitting, and ultimately enhancing the predictive capacity of GenoGraph. Importantly, the model’s robustness and consistent performance, as evidenced in the independent BCAnalyze cohort (Figure 2c), highlight its potential for practical clinical applications, offering a promising path toward more reliable genomics-based risk prediction and personalized diagnostics in oncology.

The biological relevance of the genetic variants identified by GenoGraph was validated through tissue-specific expression analyses and gene-set enrichment analysis. In doing so, it highlighted pathways like estrogen response and KRAS signaling, which have well-established links to BC, as seen in studies that have identified specific loci affecting these pathways.^77,78^ This approach not only confirms the association of these pathways with BC risk but also points towards potential molecular mechanisms that could serve as targets for therapeutic intervention.

Moreover, GenoGraph’s identification of variants predominantly located in noncoding regions, such as regulatory regions, aligns with emerging insights from GWAS studies that suggest these regions play a crucial role in cancer susceptibility. For example, regulatory variants influence gene expression through enhancer activity. This adds depth to our understanding of BC risk, moving beyond the coding regions traditionally focused on in GWAS.

### Limitations of the study

While GenoGraph demonstrates significant advancements in breast cancer risk prediction and variant interpretation, certain limitations must be acknowledged. GenoGraph’s efficacy is contingent on the quality and accuracy of input data, particularly the graph construction based on genetic similarity. Noise or inaccuracies in genotype data or the similarity metric could introduce biases, impacting predictive accuracy and biological interpretation.

Although, GenoGraph achieved promising results across the analyzed Finnish datasets, its application to multi-ethnic and or non-Finnish populations remains unexplored, limits the generalizability to border cohorts with diverse genetic backgrounds. Furthermore, the interpretation of variant-variant interactions and pathway enrichment relies on existing knowledge bases, which may be incomplete or biased, potentially limiting the comprehensiveness of the biological insights generated.

The computational complexity of GenoGraph, particularly during pretraining and fine-tuning, poses challenges for scalability to larger datasets, which may constrain its adoption in resource-limited settings. Future efforts should address these limitations by refining graph construction approaches, validating the framework in diverse populations, and optimizing computational efficiency to enhance scalability.

By integrating interpretability with robust identification of genetic risk variants, GenoGraph represents a significant advancement in genetic research, offering a scalable and versatile tool for understanding complex phenotypes. Its application extends beyond breast cancer, providing a foundation for exploring genetic interactions in other complex diseases, thereby advancing precision medicine.

## Supporting information

Supplementary results

## Data Availability

The datasets utilized in this study are not publicly available to ensure the confidentiality and privacy of patient information. Researchers interested in accessing the data may submit a formal request to the lead contact, subject to data-sharing agreements and approval. Additionally, access may be requested through the Biobank of Eastern Finland (https://ita-suomenbiopankki.fi/en/ ), which holds the datasets under appropriate ethical and institutional guidelines.

## ACKNOWLEDGMENTS

We would like to thank Jaana M. Hartikainen and Maria Tengström for providing access to the KBCP dataset, and Kaisa Luostari for valuable discussion for biological relevance interpretation and analysis. Katri Pylkäs and Robert Winqvist for providing access to OBCS dataset. We thank the Biobank of Eastern Finland for the Biobank material.

## AUTHOR CONTRIBUTIONS

Conceptualization, G.N.R., J.M.H., V.M.K., H.M., and A.M.; methodology, G.N.R., H.M., and A.M.; Investigation, G.N.R., J.M.H., H.M., and A.M.; writing—original draft, G.N.R., and H.M.; writing—review & editing, G.N.R., J.M.H., M.T., K.P., R.W., V.M.K., H.M., and A.M.; funding acquisition, V.M.K., H.M., and A.M.; resources, J.M.H., M.T., K.P., R.W., V.M.K., and A.M.; supervision, H.M., and A.M.

## METHOD DETAILS

### Variant filtering, quality checking, and preprocessing

Variant calling, filtering, and quality control for the KBCP and OBCS datasets were performed as previously described.^78^ For the BCAnalyze, we applied rigorous filtering and preprocessing methods, using PLINK.^23^ To reduce noise and redundancy, genetic variants with a call rate below 5% (missingness filter), a minor allele frequency (MAF) below 1%, and a linkage disequilibrium (LD) threshold of 𝑟² ≤ 0.2 were removed.

For all the datasets, an additive genetic model was adopted, in which genotyped variants were coded as 0 (homozygous major), 1 (heterozygous), or 2 (homozygous minor), representing the count of minor alleles per individual.^9^ We normalized the obtained genotype matrix 𝑋 ∈ ℝ^n×m^, where 𝑛 and 𝑚 represent the number of individuals and SNPs, respectively, as

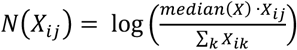

where 𝑚𝑒𝑑𝑖𝑎𝑛(𝑋) indicates the median value of the genotype data across all SNPs, providing a central tendency measure for rescaling. The genotype matrix 𝑋 was transformed using the natural logarithm to smooth and rescale the data, thereby reducing the influence of extreme values and enhancing interpretability (Figure S1). Pairwise correlation filtering was subsequently applied to exclude variants exhibiting high correlation (𝑟 ≥ 0.6).

### Graph representation

The resulting genotype matrix 𝑋 ∈ ℝ^n×d^, where 𝑑 represents the number of filtered variants, was then used to construct the graph 𝒢, which represents the relation between the individuals with genetic similarity, based on Wang, B., et al^24^. Initially, the genetic similarity between any two individuals 𝑥 and 𝑦 is quantified using the weighted Hamming distance^79^ 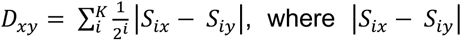 represents the absolute difference between 𝑥 and 𝑦 individuals for SNP 𝑖. 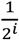 assigns a weight to the absolute difference. If 𝐷_xy_ < 0.5, then there exists an edge connection between the nodes (individuals) 𝑥 and 𝑦. The weighted 𝒢 effectively captures the complex interactions among individuals (nodes) and encoded genetic variants (node features), thereby enhancing the ability to predict individual genetic risk models with improved precision and biological insights.

### Data splitting protocol

We employed a rigorous and systematic approach for dataset partitioning, ensuring robust model development and evaluation across all three datasets. Specifically, each dataset was stratified and divided into a training set comprising 70% of the data and an independent test set comprising the remaining 30%. The training set was used primarily for model development, including both learning and hyperparameter tuning procedures.

To achieve optimal model tuning while mitigating overfitting, we implemented a nested 5-fold cross- validation approach. In this configuration, the inner loop of the nested cross-validation^80^ consisted of one- fold, which was dedicated to hyperparameter tuning using Optuna.^81^ The outer loop comprised five folds, wherein four folds were employed for model training, and the remaining fold was used to monitor the development process and assess interim performance. This nested cross-validation strategy enables reliable selection of hyperparameters while preserving a rigorous distinction between training, validation, and evaluation phases.

The independent test set, which was never used during training or hyperparameter optimization, was used for final performance evaluation, allowing an unbiased estimation of the model’s predictive capability. To further quantify the uncertainty in our performance metrics, we employed a bootstrapping^82^ technique with 500 resamples to compute 95% confidence intervals for key evaluation metrics, providing a more comprehensive understanding of the variability and robustness of the model’s predictions.

### GenoGraph pre-training details

Figure 1 illustrates the proposed sparse graph variational autoencoder (SGVAE)-based contrastive learning framework as a pre-training model, designed for robust variant representation learning. We employed infoNCE loss as contrastive loss to minimize the distance between the positive pairs (same nodes from 𝒢_1_and 𝒢_2_), while maximizing the distance between negative pairs.^83, 84^ Furthermore, we employed a self- attention^85^ module to capture the intricate complex genetic relation within the individual using the latent representations 𝑍 ∈ ℝ^n×5^^12^. The contrastive loss is supervised with the class labels (case or control) provided within the KBCP, OBCS, and BCAnalyze datasets.^20,21^

We combined the training sets of KBCP and OBCS datasets to pre-train the GenoGraph framework using the combination of contrastive, reconstruction, and classification loss functions, defined as

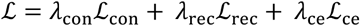

For contrastive loss ℒ_con_, specifically, we employed infoNCE loss^84^, which is designed to pull positive pairs (similar nodes of 𝒢_%_ and 𝒢_&_) closer in the embedding space while pushing negative pairs apart,

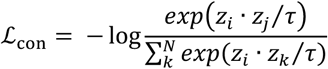

where, 𝑧_i_ and 𝑧_j_are the embeddings of positive pairs, 𝑧_k_ represents embeddings from the negative sample set, 𝜏 is a temperate parameter that scales the distribution of the distance, and 𝑁 is the total number of negative samples plus one positive sample. For reconstruction loss ℒ_rec_ we employed the MSE loss with KL-divergence^86^ to account for both the accuracy of the reconstruction and the alignment of the learned latent distribution with the prior, defined as

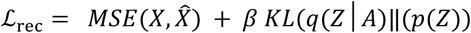

where 𝑋 and 𝑋 are the original and reconstructed graphs, respectively. 𝑞(𝑍│𝐴) is the posterior distribution, 𝑝(𝑍) is the prior distribution, and 𝛽 is the KL-divergence controlling factor. For the classification loss ℒ_ce_, we employed binary cross-entropy loss. 𝜆_con_, 𝜆_rec_, 𝑎𝑛𝑑 𝜆_ce_are the weight coefficents to balance the combined loss. In our experiments, we defined 𝜆_con_ = 0.50, 𝜆_rec_ = 0.25, 𝑎𝑛𝑑 𝜆_ce_ = 0.25 based on hyper-parameter tuning.

#### Fine-tuning GenoGraph for downstream analysis

We fine-tuned the pretrained GenoGraph framework, specifically using the trained weights of SGVAE encoder to extract the variant embeddings 𝑍 ∈ ℝ^n×1^^52^, subsequently given as input for the self-attention, and finally given to the SoftPlus^87^ as the classification layer. In this study, we considered the case–control node classification as one of the downstream analysis tasks.

For the case–-control node classification, we employed the focal Tversky loss^88^.

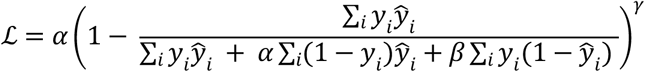

where, 𝑦_i_and 𝑦i represent the actual and predicted labels for the node 𝑖 in 𝒢. 𝛼, and 𝛽 are the parameters that control the penalty for the false positive and false negatives, respectively. 𝛾 adjusts the contribution of hard-to-classify nodes in 𝒢. In this study, we used 𝛼 = 0.3, 𝛽 = 0.7, and 𝛾 = 1 as the optimal values, as suggested by Abraham, N. and Khan, N.M.,^88^.

#### Benchmarking against ML, DL and Graph based GWAS approaches

We compared our GenoGraph to:

**Chi-Square Test** evaluates the independence between categorical variables, such as genetic variants (SNPs) and phenotypic outcomes. It identifies SNPs with significant deviations from the expected distribution under the null hypothesis, making it particularly effective for categorical trait analysis in GWAS.

**ANOVA (Analysis of Variance)** assesses differences in means across genotype groups (e.g., homozygous dominant, heterozygous, and homozygous recessive) for continuous traits. By quantifying the variance explained by genotypes, ANOVA identifies loci significantly associated with quantitative phenotypes, aiding GWAS investigations.

**Mutual Information (MI)** measures the dependency between genetic variants and traits, capturing both linear and non-linear relationships. According to Beraha et al.,^34^ MI provides a robust framework for FS, offering theoretical guarantees and adaptability to GWAS data’s complexity. It ensures that relevant SNPs with intricate dependency structures are retained.

**Random Forest (RF)** an ensemble-based ML method that computes feature importance based on the decrease in impurity (e.g., Gini importance) when a SNP is used for a split. While effective for ranking SNPs, RF can favor variants with high minor allele frequencies, requiring careful interpretation in GWAS studies.^35^

**ElasticNet** a linear model with combined L1 (Lasso) and L2 (Ridge) penalties, is well-suited for GWAS due to its ability to manage high-dimensional genomic data and feature collinearity. It jointly selects correlated SNPs and provides stable models even with limited sample sizes.^36^

**HSIC Lasso** a kernel-based method that leverages the Hilbert-Schmidt Independence Criterion (HSIC) to capture complex, non-linear dependencies between SNPs and traits. This approach is particularly effective for high-dimensional data, making it a powerful tool for GWAS where non-linear effects are common.^37^

**LASSONet** integrates Lasso regularization within a neural network framework. It offers scalable FS and models SNP interactions while ensuring sparsity. This dual capability makes it suitable for identifying influential SNPs in GWAS datasets with intricate dependency structures.^38^

**Deep Neural Pursuit (DNP)** introduced by Liu et al.,^39^ is designed for high-dimensional, low-sample-size datasets typical of GWAS. It uses deep neural networks (DNNs) to iteratively select SNPs by learning their influence on phenotypic traits, providing scalability and robustness against overfitting.

**Deep Feature Selection (DFS)** Li et al.,^40^ developed DFS to incorporate sparsity-inducing penalties directly into DL models. This method identifies relevant SNPs during model training, ensuring that the FS process is integrated with predictive modeling, enhancing interpretability in GWAS.

**Deep Feature Screening (DeepFS)** Li et al.,^41^ proposed DeepFS for ultra-high-dimensional data. This method optimizes predictive performance and sparsity simultaneously using DNNs. Its suitability for GWAS lies in its ability to efficiently handle vast numbers of SNPs while identifying significant variants.

**GRACES** Chen et al.,^42^ introduced GRACES, a graph convolutional network-based method that leverages feature relationships (e.g., linkage disequilibrium) to guide selection. This approach is particularly relevant to GWAS, as it incorporates biological relevance by modeling SNP interactions within a graph structure.

#### Functional variant annotation and gene mapping

We employed SNPxplorer^43^, a web-based tool designed for comprehensive variant annotation and gene mapping. SNPxplorer annotates genetic variants by integrating data from various bioinformatics resources to provide detailed functional insights. Reference databases such as Ensembl, dbSNP, and the 1000 Genomes Project were used to obtain precise positional and contextual information. Functional consequences of variants were determined using tools such as the Ensembl Variant Effect Predictor (VEP), which classified variants into synonymous, missense, regulatory, and other categories. SNPxplorer incorporates regulatory annotations from sources such as RegulomeDB and ENCODE to identify the effects of variants in non-coding regions. LD data were also included to understand broader genetic contexts and relationships, facilitating the interpretation of variant impacts within gene networks.

For gene mapping, SNPxplorer integrates expression quantitative trait loci (eQTL) data from the Genotype- Tissue Expression (GTEx) version 8 database to establish variant–gene relationships.^89^ In this analysis, eQTL data from breast tissue was specifically used to map variants to genes of potential regulatory relevance. When eQTL data were unavailable, a positional approach was employed, mapping variants to the nearest gene within a 50 kb range.

Furthermore, SNPxplorer computed additional annotations for significant variants, including Combined Annotation Dependent Depletion (CADD) scores,^90^ PolyPhen scores,^45^ and SIFT scores,^44^ which provided insights into the deleteriousness and potential functional impact of the variants. These comprehensive annotations are crucial for understanding the biological relevance of identified SNPs and their contribution to phenotypic traits or diseases.

#### Gene-set enrichment analysis

We employed the gene set enrichment analysis (GSEA) algorithm using the Python package GSEApy to determine the biological significance of variants identified by the GenoGraph framework.^46,91^ GSEA ranks genes based on their association with a phenotype of interest and calculates an enrichment score (ES) for each pathway using a weighted Kolmogorov-Smirnov-like statistic.^92^ A high ES indicates that a pathway’s genes are predominantly found at either end of the ranked list, reflecting significant enrichment.

A total of 2,500 variants identified by GenoGraph and their genes (mapped using SNPxplorer) were analyzed using GSEA with two major gene sets: MSigDB Hallmark 2020^93^ and KEGG pathways^94^. MSigDB Hallmark includes 50 gene sets representing major biological processes, while KEGG provides a comprehensive representation of molecular networks. To ensure robustness, a Bonferroni correction was applied to control for multiple comparisons across the 2,124 gene sets, thereby minimizing false positives.

#### Tissue-type gene expression

To evaluate the biological significance of specific tissue types in relation to our analysis, we analyzed tissue- type gene expression in detail using FUMA (Functional Mapping and Annotation),^95^ specifically employing the *gene2Function* module. This analysis aimed to determine whether differential gene expression data across a diverse range of tissue types could help elucidate the biological relevance of variants identified by the GenoGraph framework.

We assessed gene expression data for 30 broad tissue types and 53 specific tissues, with all tissues sourced from the Genotype-Tissue Expression (GTEx) V8 database.^89^ Gene expression values were transformed using a log2 scale with an added pseudo count of 1 to handle zero expression values, ensuring data consistency and preventing computational issues due to zero counts. Additionally, the data were winsorized at the 50th percentile to mitigate the influence of extreme outliers, which can otherwise bias downstream analyses. The average expression value for each tissue type was subsequently calculated and used as a representative measure in further evaluations.

Multiple-testing correction was performed using the Bonferroni method for the two sets of analyses (30 broad tissue types and 53 specific tissues). This stringent correction helped control the false discovery rate, ensuring that any significant association identified between tissue-specific differential expression levels and GenoGraph-identified variants was robust and reliable. This allowed us to systematically assess the contribution of these tissue types to the biological processes influenced by the identified genetic variants, providing a clearer understanding of how tissue-specific expression patterns are linked to disease- associated genetic alterations.

## SUPPLEMENTAL INFORMATION

**Document S1. Figures S1–S3** (this is the main PDF)

**Document S2. Tables 1-11**

## REFERENCES

1. Sun, Y.S., Zhao, Z., Yang, Z.N., Xu, F., Lu, H.J., Zhu, Z.Y., Shi, W., Jiang, J., Yao, P.P. and Zhu, H.P., 2017. Risk factors and preventions of breast cancer. International journal of biological sciences, 13(11), p.1387.

2. Obeagu, E.I. and Obeagu, G.U., 2024. Breast cancer: A review of risk factors and diagnosis. Medicine, 103(3), p.e36905.

3. McTiernan, A., Kuniyuki, A., Yasui, Y., Bowen, D., Burke, W., Culver, J.B., Anderson, R. and Durfy, S., 2001. Comparisons of two breast cancer risk estimates in women with a family history of breast cancer. Cancer Epidemiology Biomarkers & Prevention, 10(4), pp.333–338.

4. Fackenthal, J.D. and Olopade, O.I., 2007. Breast cancer risk associated with BRCA1 and BRCA2 in diverse populations. Nature Reviews Cancer, 7(12), pp.937–948.

5. Erkko, H., Xia, B., Nikkilä, J., Schleutker, J., Syrjäkoski, K., Mannermaa, A., Kallioniemi, A., Pylkäs, K., Karppinen, S.M., Rapakko, K. and Miron, A., 2007. A recurrent mutation in PALB2 in Finnish cancer families. Nature, 446(7133), pp.316–319.

6. Hollestelle, A., Wasielewski, M., Martens, J.W. and Schutte, M., 2010. Discovering moderate-risk breast cancer susceptibility genes. Current opinion in genetics & development, 20(3), pp.268–276.

7. Mahdi, K.M., Nassiri, M.R. and Nasiri, K., 2013. Hereditary genes and SNPs associated with breast cancer. Asian Pacific Journal of Cancer Prevention, 14(6), pp.3403–3409.

8. Mars, N., Widén, E., Kerminen, S., Meretoja, T., Pirinen, M., della Briotta Parolo, P., Palta, P., Palotie, A., Kaprio, J. and Joensuu, H., 2020. The role of polygenic risk and susceptibility genes in breast cancer over the course of life. Nature communications, 11(1), p.6383.

9. Hayes, B., 2013. Overview of statistical methods for genome-wide association studies (GWAS). Genome-wide association studies and genomic prediction, pp.149–169.

10. Sollis, E., Mosaku, A., Abid, A., Buniello, A., Cerezo, M., Gil, L., Groza, T., Güneş, O., Hall, P., Hayhurst, J. and Ibrahim, A., 2023. The NHGRI-EBI GWAS Catalog: knowledgebase and deposition resource. Nucleic acids research, 51(D1), pp.D977–D985.

11. Behravan, H., Hartikainen, J.M., Tengström, M., Kosma, V.M. and Mannermaa, A., 2020. Predicting breast cancer risk using interacting genetic and demographic factors and machine learning. Scientific reports, 10(1), p.11044.

12. Behravan, H., Hartikainen, J.M., Tengström, M., Pylkäs, K., Winqvist, R., Kosma, V.M. and Mannermaa, A., 2018. Machine learning identifies interacting genetic variants contributing to breast cancer risk: A case study in Finnish cases and controls. Scientific reports, 8(1), p.13149.

13. Sigala, R.E., Lagou, V., Shmeliov, A., Atito, S., Kouchaki, S., Awais, M., Prokopenko, I., Mahdi, A. and Demirkan, A., 2023. Machine Learning to Advance Human Genome-Wide Association Studies. Genes, 15(1), p.34.

14. Lever, J., Krzywinski, M. and Altman, N., 2016. Points of significance: model selection and overfitting. Nature methods, 13(9), pp.703–705.

15. Pudjihartono, N., Fadason, T., Kempa-Liehr, A.W. and O’Sullivan, J.M., 2022. A review of feature selection methods for machine learning-based disease risk prediction. Frontiers in Bioinformatics, 2, p.927312.

16. Nguyen, T.T., Huang, J.Z., Wu, Q., Nguyen, T.T. and Li, M.J., 2015, December. Genome-wide association data classification and SNPs selection using two-stage quality-based Random Forests. In BMC genomics (Vol. 16, pp. 1–11). BioMed Central.

17. Jo, T., Nho, K., Bice, P., Saykin, A.J. and Alzheimer’s Disease Neuroimaging Initiative, 2022. Deep learning-based identification of genetic variants: application to Alzheimer’s disease classification. Briefings in Bioinformatics, 23(2), p.bbac022.

18. Li, Y., Wen, J., Li, G., Chen, J., Sun, Q., Liu, W., Guan, W., Lai, B., Szatkiewicz, J., He, X. and Sullivan, P., 2023. DeepGWAS: Enhance GWAS Signals for Neuropsychiatric Disorders via Deep Neural Network. Research Square.

19. Arloth, J., Eraslan, G., Andlauer, T.F., Martins, J., Iurato, S., Kühnel, B., Waldenberger, M., Frank, J., Gold, R., Hemmer, B. and Luessi, F., 2020. DeepWAS: Multivariate genotype-phenotype associations by directly integrating regulatory information using deep learning. PLoS computational biology, 16(2), p.e1007616.

20. Tan, Z., Ding, K., Guo, R. and Liu, H., 2022, September. Supervised graph contrastive learning for few-shot node classification. In Joint European Conference on Machine Learning and Knowledge Discovery in Databases (pp. 394-411). Cham: Springer International Publishing.

21. Ji, J., Jia, H., Ren, Y. and Lei, M., 2023. Supervised contrastive learning with structure inference for graph classification. IEEE Transactions on Network Science and Engineering, 10(3), pp.1684–1695.

22. Ying, Z., Bourgeois, D., You, J., Zitnik, M. and Leskovec, J., 2019. Gnnexplainer: Generating explanations for graph neural networks. Advances in neural information processing systems, 32.

23. Purcell, S., Neale, B., Todd-Brown, K., Thomas, L., Ferreira, M.A., Bender, D., Maller, J., Sklar, P., De Bakker, P.I., Daly, M.J. and Sham, P.C., 2007. PLINK: a tool set for whole-genome association and population-based linkage analyses. The American journal of human genetics, 81(3), pp.559–575.

24. Wang, B., Mezlini, A.M., Demir, F., Fiume, M., Tu, Z., Brudno, M., Haibe-Kains, B. and Goldenberg, A., 2014. Similarity network fusion for aggregating data types on a genomic scale. Nature methods, 11(3), pp.333–337.

25. Yu, J., Yin, H., Xia, X., Chen, T., Cui, L. and Nguyen, Q.V.H., 2022, July. Are graph augmentations necessary? simple graph contrastive learning for recommendation. In Proceedings of the 45th international ACM SIGIR conference on research and development in information retrieval (pp. 1294-1303).

26. Kipf, T.N. and Welling, M., 2016. Semi-supervised classification with graph convolutional networks. arXiv preprint arXiv:1609.02907.

27. Veličković, P., Cucurull, G., Casanova, A., Romero, A., Lio, P. and Bengio, Y., 2017. Graph attention networks. arXiv preprint arXiv:1710.10903.

28. Hamilton, W., Ying, Z. and Leskovec, J., 2017. Inductive representation learning on large graphs. Advances in neural information processing systems, 30.

29. Xu, J., Sun, X., Zhang, Z., Zhao, G. and Lin, J., 2019. Understanding and improving layer normalization. Advances in neural information processing systems, 32.

30. Gal, Y. and Ghahramani, Z., 2015. Dropout as a Bayesian approximation. arXiv preprint arXiv:1506.02157.

31. Geng, W., An, J., Dong, K., Zhang, H., Zhang, X., Liu, Y., Xu, R., Liu, Y., Huang, X., Song, H. and Yan, W., 2024. ZNF8 Orchestrates with Smad3 to Promote Lung Metastasis by Recruiting SMYD3 in Breast Cancer. Advanced Science, p.2404904.

32. Ghaleb, A.M. and Yang, V.W., 2017. Krüppel-like factor 4 (KLF4): What we currently know. Gene, 611, pp.27–37.

33. Zhang, L. and Long, X., 2015. Association of three SNPs in TOX3 and breast cancer risk: Evidence from 97275 cases and 128686 controls. Scientific reports, 5(1), p.12773.

34. Beraha, M., Metelli, A.M., Papini, M., Tirinzoni, A. and Restelli, M., 2019, July. Feature selection via mutual information: New theoretical insights. In 2019 international joint conference on neural networks (IJCNN) (pp. 1-9). IEEE.

35. Boulesteix, A.L., Bender, A., Lorenzo Bermejo, J. and Strobl, C., 2012. Random forest Gini importance favours SNPs with large minor allele frequency: impact, sources and recommendations. Briefings in Bioinformatics, 13(3), pp.292–304.

36. Cho, S., Kim, K., Kim, Y.J., Lee, J.K., Cho, Y.S., Lee, J.Y., Han, B.G., Kim, H., Ott, J. and Park, T., 2010. Joint identification of multiple genetic variants via elastic-net variable selection in a genome-wide association analysis. Annals of human genetics, 74(5), pp.416–428.

37. Yamada, M., Jitkrittum, W., Sigal, L., Xing, E.P. and Sugiyama, M., 2014. High-dimensional feature selection by feature-wise kernelized lasso. Neural computation, 26(1), pp.185–207.

38. Lemhadri, I., Ruan, F., Abraham, L. and Tibshirani, R., 2021. Lassonet: A neural network with feature sparsity. Journal of Machine Learning Research, 22(127), pp.1–29.

39. Liu, B., Wei, Y., Zhang, Y. and Yang, Q., 2017, August. Deep Neural Networks for High Dimension, Low Sample Size Data. In IJCAI (Vol. 2017, pp. 2287–2293).

40. Li, Y., Chen, C.Y. and Wasserman, W.W., 2016. Deep feature selection: theory and application to identify enhancers and promoters. Journal of Computational Biology, 23(5), pp.322–336.

41. Li, K., Wang, F., Yang, L. and Liu, R., 2023. Deep feature screening: Feature selection for ultra high-dimensional data via deep neural networks. Neurocomputing, 538, p.126186.

42. Chen, C., Weiss, S.T. and Liu, Y.Y., 2023. Graph convolutional network-based feature selection for high-dimensional and low-sample size data. Bioinformatics, 39(4), p.btad135.

43. Tesi, N., Van Der Lee, S., Hulsman, M., Holstege, H. and Reinders, M.J., 2021. snpXplorer: a web application to explore human SNP-associations and annotate SNP-sets. Nucleic acids research, 49(W1), pp.W603–W612.

44. Kumar, P., Henikoff, S. and Ng, P.C., 2009. Predicting the effects of coding non-synonymous variants on protein function using the SIFT algorithm. Nature protocols, 4(7), pp.1073–1081.

45. Adzhubei, I., Jordan, D.M. and Sunyaev, S.R., 2013. Predicting functional effect of human missense mutations using PolyPhen-2. Current protocols in human genetics, 76(1), pp.7–20.

46. Fang, Z., Liu, X. and Peltz, G., 2023. GSEApy: a comprehensive package for performing gene set enrichment analysis in Python. Bioinformatics, 39(1), p.btac757.

47. Abedin, M.J., Wang, D., McDonnell, M.A., Lehmann, U. and Kelekar, A., 2007. Autophagy delays apoptotic death in breast cancer cells following DNA damage. Cell Death & Differentiation, 14(3), pp.500–510.

48. Cattoretti, G., Rilke, F., Andreola, S., D’Amato, L. and Delia, D., 1988. p53 expression in breast cancer. International Journal of Cancer, 41(2), pp.178–183.

49. Wooster, R., Bignell, G., Lancaster, J., Swift, S., Seal, S., Mangion, J., Collins, N., Gregory, S., Gumbs, C., Micklem, G. and Barfoot, R., 1995. Identification of the breast cancer susceptibility gene BRCA2. Nature, 378(6559), pp.789–792.

50. Jeselsohn, R., Buchwalter, G., De Angelis, C., Brown, M. and Schiff, R., 2015. ESR1 mutations— a mechanism for acquired endocrine resistance in breast cancer. Nature reviews Clinical oncology, 12(10), pp.573–583.

51. Cizkova, M., Vacher, S., Meseure, D., Trassard, M., Susini, A., Mlcuchova, D., Callens, C., Rouleau, E., Spyratos, F., Lidereau, R. and Bièche, I., 2013. PIK3R1 underexpression is an independent prognostic marker in breast cancer. BMC cancer, 13, pp.1–15.

52. Strocchi, S., Reggiani, F., Gobbi, G., Ciarrocchi, A. and Sancisi, V., 2022. The multifaceted role of EGLN family prolyl hydroxylases in cancer: going beyond HIF regulation. Oncogene, 41(29), pp.3665–3679.

53. Bjarnadottir, O., Feldt, M., Inasu, M., Bendahl, P.O., Elebro, K., Kimbung, S. and Borgquist, S., 2020. Statin use, HMGCR expression, and breast cancer survival–The Malmö Diet and Cancer Study. Scientific reports, 10(1), p.558.

54. Wouters, B.G. and Koritzinsky, M., 2008. Hypoxia signalling through mTOR and the unfolded protein response in cancer. Nature Reviews Cancer, 8(11), pp.851–864.

55. Petroulakis, E., Mamane, Y., Le Bacquer, O., Shahbazian, D. and Sonenberg, N., 2006. mTOR signaling: implications for cancer and anticancer therapy. British journal of cancer, 94(2), pp.195–199.

56. Zhang, L. and Long, X., 2015. Association of three SNPs in TOX3 and breast cancer risk: Evidence from 97275 cases and 128686 controls. Scientific reports, 5(1), p.12773.

57. Müller, A., Homey, B., Soto, H., Ge, N., Catron, D., Buchanan, M.E., McClanahan, T., Murphy, E., Yuan, W., Wagner, S.N. and Barrera, J.L., 2001. Involvement of chemokine receptors in breast cancer metastasis. nature, 410(6824), pp.50–56.

58. Liang, Y., Han, H., Liu, L., Duan, Y., Yang, X., Ma, C., Zhu, Y., Han, J., Li, X. and Chen, Y., 2018. CD36 plays a critical role in proliferation, migration and tamoxifen-inhibited growth of ER-positive breast cancer cells. Oncogenesis, 7(12), p.98.

59. Feng, W.W., Wilkins, O., Bang, S., Ung, M., Li, J., An, J., Del Genio, C., Canfield, K., DiRenzo, J., Wells, W. and Gaur, A., 2019. CD36-mediated metabolic rewiring of breast cancer cells promotes resistance to HER2-targeted therapies. Cell reports, 29(11), pp.3405–3420.

60. Cheng, Q., Jabbari, K., Winkelmaier, G., Andersen, C., Yaswen, P., Khoshdeli, M. and Parvin, B., 2020. Overexpression of CD36 in mammary fibroblasts suppresses colony growth in breast cancer cell lines. Biochemical and biophysical research communications, 526(1), pp.41–47.

61. Ligorio, F., Di Cosimo, S., Verderio, P., Ciniselli, C.M., Pizzamiglio, S., Castagnoli, L., Dugo, M., Galbardi, B., Salgado, R., Loi, S. and Michiels, S., 2022. Predictive role of CD36 expression in HER2-positive breast cancer patients receiving neoadjuvant trastuzumab. JNCI: Journal of the National Cancer Institute, 114(12), pp.1720–1727.

62. Linardou, H., Kalogeras, K.T., Kronenwett, R., Alexopoulou, Z., Wirtz, R.M., Zagouri, F., Scopa, C.D., Gogas, H., Petraki, K., Christodoulou, C. and Pavlakis, K., 2015. Prognostic significance of VEGFC and VEGFR1 mRNA expression according to HER2 status in breast cancer: a study of primary tumors from patients with high-risk early breast cancer participating in a randomized Hellenic Cooperative Oncology Group Trial. Anticancer research, 35(7), pp.4023–4036.

63. Banerjee, K., Kerzel, T., Bekkhus, T., de Souza Ferreira, S., Wallmann, T., Wallerius, M., Landwehr, L.S., Agardy, D.A., Schauer, N., Malmerfeldt, A. and Bergh, J., 2023. VEGF-C-expressing TAMs rewire the metastatic fate of breast cancer cells. Cell Reports, 42(12).

64. Pal, S.K., Nguyen, C.T.K., Morita, K.I., Miki, Y., Kayamori, K., Yamaguchi, A. and Sakamoto, K., 2016. THBS 1 is induced by TGFB 1 in the cancer stroma and promotes invasion of oral squamous cell carcinoma. Journal of oral pathology & medicine, 45(10), pp.730–739.

65. Li, Y., Qin, J., Chen, G., Wu, W. and Sun, X., 2024. Plasma THBS1 as a predictive biomarker for poor prognosis and brain metastasis in patients with HER2-enriched breast cancer. International Journal of Clinical Oncology, 29(4), pp.427–441.

66. Zhang, C., Hu, C., Su, K., Wang, K., Du, X., Xing, B. and Liu, X., 2021. The integrative analysis of thrombospondin family genes in Pan-cancer reveals that THBS2 facilitates gastrointestinal cancer metastasis. Journal of oncology, 2021(1), p.4405491.

67. Tsuchiya, Y., Nakajima, M., Kyo, S., Kanaya, T., Inoue, M. and Yokoi, T., 2004. Human CYP1B1 is regulated by estradiol via estrogen receptor. Cancer research, 64(9), pp.3119–3125.

68. De Vivo, I., Hankinson, S.E., Li, L., Colditz, G.A. and Hunter, D.J., 2002. Association of CYP1B1 polymorphisms and breast cancer risk. Cancer Epidemiology Biomarkers & Prevention, 11(5), pp.489–492.

69. Economopoulos, K.P. and Sergentanis, T.N., 2010. Three polymorphisms in cytochrome P450 1B1 (CYP1B1) gene and breast cancer risk: a meta-analysis. Breast cancer research and treatment, 122, pp.545–551.

70. Wang, F., Jiang, L., Yu, X., Li, M., Wu, G., Yu, Z., Zhou, K., Chu, H. and Zhai, H., 2015. Association between TCF7L2 polymorphisms and breast cancer susceptibility: a meta-analysis. International Journal of Clinical and Experimental Medicine, 8(6), p.9355.

71. Zheng, A., Song, X., Zhang, L., Zhao, L., Mao, X., Wei, M. and Jin, F., 2019. Long non-coding RNA LUCAT1/miR-5582-3p/TCF7L2 axis regulates breast cancer stemness via Wnt/β-catenin pathway. Journal of Experimental & Clinical Cancer Research, 38, pp.1–14.

72. Burwinkel, B., Shanmugam, K.S., Hemminki, K., Meindl, A., Schmutzler, R.K., Sutter, C., Wappenschmidt, B., Kiechle, M., Bartram, C.R. and Frank, B., 2006. Transcription factor 7-like 2 (TCF7L2) variant is associated with familial breast cancer risk: a case-control study. BMC cancer, 6, pp.1–4.

73. Shi, Q., Xue, C., Zeng, Y., Yuan, X., Chu, Q., Jiang, S., Wang, J., Zhang, Y., Zhu, D. and Li, L., 2024. Notch signaling pathway in cancer: from mechanistic insights to targeted therapies. Signal Transduction and Targeted Therapy, 9(1), p.128.

74. Mancino, M., Ametller, E., Gascón, P. and Almendro, V., 2011. The neuronal influence on tumor progression. Biochimica et Biophysica Acta (BBA)-Reviews on Cancer, 1816(2), pp.105–118.

75. Shi, Y., Luo, J., Wang, X., Zhang, Y., Zhu, H., Su, D., Yu, W. and Tian, J., 2022. Emerging trends on the correlation between neurotransmitters and tumor progression in the last 20 years: a bibliometric analysis via CiteSpace. Frontiers in Oncology, 12, p.800499.

76. Strocchi, S., Reggiani, F., Gobbi, G., Ciarrocchi, A. and Sancisi, V., 2022. The multifaceted role of EGLN family prolyl hydroxylases in cancer: going beyond HIF regulation. Oncogene, 41(29), pp.3665–3679.

77. Jia, G., Ping, J., Guo, X., Yang, Y., Tao, R., Li, B., Ambs, S., Barnard, M.E., Chen, Y., Garcia-Closas, M. and Gu, J., 2024. Genome-wide association analyses of breast cancer in women of African ancestry identify new susceptibility loci and improve risk prediction. Nature genetics, pp.1–8.

78. Michailidou, K., Beesley, J., Lindstrom, S., Canisius, S., Dennis, J., Lush, M.J., Maranian, M.J., Bolla, M.K., Wang, Q., Shah, M. and Perkins, B.J., 2015. Genome-wide association analysis of more than 120,000 individuals identifies 15 new susceptibility loci for breast cancer. Nature genetics, 47(4),

79. Bookstein, A., Kulyukin, V.A. and Raita, T., 2002. Generalized hamming distance. Information Retrieval, 5, pp.353–375.

80. Wainer, J. and Cawley, G., 2021. Nested cross-validation when selecting classifiers is overzealous for most practical applications. Expert Systems with Applications, 182, p.115222.

81. Akiba, T., Sano, S., Yanase, T., Ohta, T. and Koyama, M., 2019, July. Optuna: A next-generation hyperparameter optimization framework. In Proceedings of the 25th ACM SIGKDD international conference on knowledge discovery & data mining (pp. 2623-2631).

82. Diciccio, T.J. and Romano, J.P., 1988. A review of bootstrap confidence intervals. Journal of the Royal Statistical Society Series B: Statistical Methodology, 50(3), pp.338–354.

83. Chen, T., Kornblith, S., Norouzi, M. and Hinton, G., 2020, November. A simple framework for contrastive learning of visual representations. In International conference on machine learning (pp. 1597-1607). PMLR.

84. Rusak, E., Reizinger, P., Juhos, A., Bringmann, O., Zimmermann, R.S. and Brendel, W., 2024. InfoNCE: Identifying the Gap Between Theory and Practice. arXiv preprint arXiv:2407.00143.

85. Vaswani, A., 2017. Attention is all you need. Advances in Neural Information Processing Systems.

86. Feng, L., Wang, H., Jin, B., Li, H., Xue, M. and Wang, L., 2018. Learning a distance metric by balancing kl-divergence for imbalanced datasets. IEEE Transactions on Systems, Man, and Cybernetics: Systems, 49(12), pp.2384–2395.

87. Zheng, H., Yang, Z., Liu, W., Liang, J. and Li, Y., 2015, July. Improving deep neural networks using softplus units. In 2015 International joint conference on neural networks (IJCNN) (pp. 1-4). IEEE.

88. Abraham, N. and Khan, N.M., 2019, April. A novel focal tversky loss function with improved attention u-net for lesion segmentation. In 2019 IEEE 16th international symposium on biomedical imaging (ISBI 2019) (pp. 683-687). IEEE.

89. Lonsdale, J., Thomas, J., Salvatore, M., Phillips, R., Lo, E., Shad, S., Hasz, R., Walters, G., Garcia, F., Young, N. and Foster, B., 2013. The genotype-tissue expression (GTEx) project. Nature genetics, 45(6), pp.580–585.

90. Kircher, M., Witten, D.M., Jain, P., O’roak, B.J., Cooper, G.M. and Shendure, J., 2014. A general framework for estimating the relative pathogenicity of human genetic variants. Nature genetics, 46(3), pp.310–315.

91. Merico, D., Isserlin, R., Stueker, O., Emili, A. and Bader, G.D., 2010. Enrichment map: a network-based method for gene-set enrichment visualization and interpretation. PloS one, 5(11), p.e13984.

92. Charmpi, K. and Ycart, B., 2015. Weighted Kolmogorov Smirnov testing: an alternative for gene set enrichment analysis. Statistical applications in genetics and molecular biology, 14(3), pp.279–293.

93. Liberzon, A., Birger, C., Thorvaldsdóttir, H., Ghandi, M., Mesirov, J.P. and Tamayo, P., 2015. The molecular signatures database hallmark gene set collection. Cell systems, 1(6), pp.417–425.

94. Kanehisa, M., 2002, November. The KEGG database. In ‘In silico’simulation of biological processes: Novartis Foundation Symposium 247 (Vol. 247, pp. 91-103). Chichester, UK: John Wiley & Sons, Ltd.

95. Watanabe, K., Taskesen, E., Van Bochoven, A. and Posthuma, D., 2017. Functional mapping and annotation of genetic associations with FUMA. Nature communications, 8(1), p.1826

