## Supplementary figures for "An Interpretable Sparse Graph Contrastive Learning Approach for Identifying Breast Cancer Risk Variants"

### Supplementary material

**Fig. S1: Variant pre-processing pipeline using PLINK and patient similarity matrix representation.**

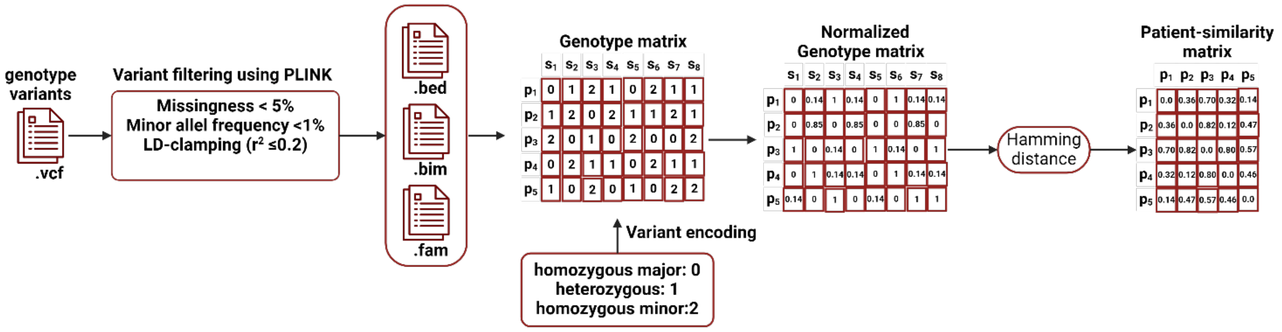


**Fig. S2: Comparison of feature selection methods for high dimensional and low sample size data.**

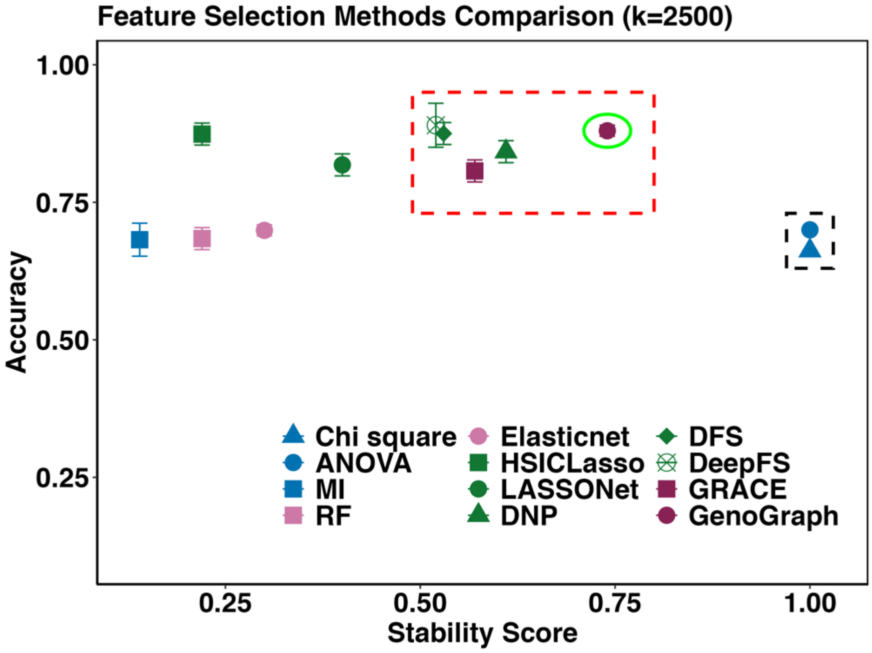

**Fig. S3: Manhattan plot of the GenoGraph identified optimal subset of SNPs.**

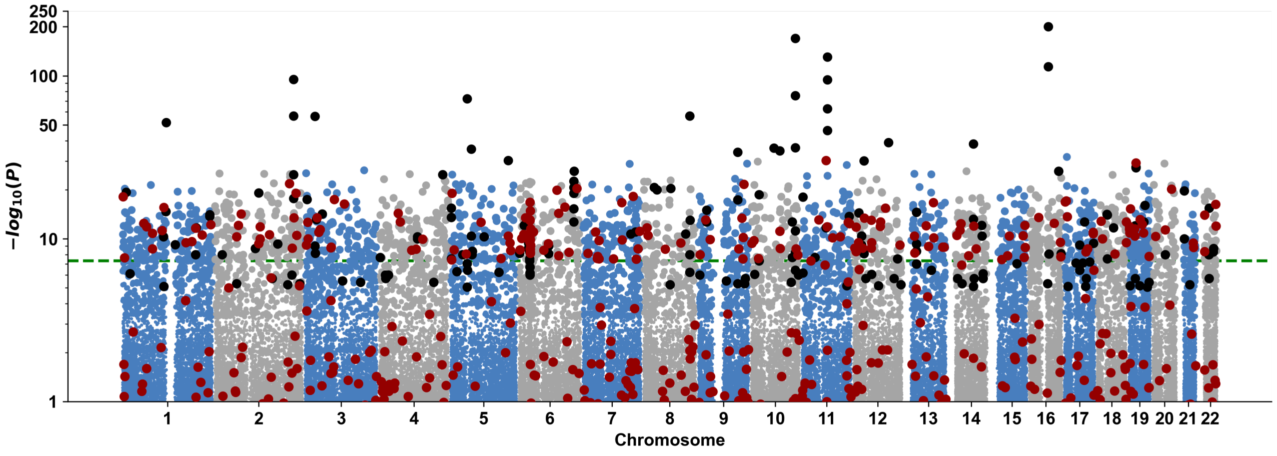
